# The Effect of Conflict on Healthcare Workers in Syria: Results of a Qualitative Survey

**DOI:** 10.1101/2021.09.27.21264178

**Authors:** Sarah Abdelrahman, Rohini J. Haar

## Abstract

The purpose of this study is to understand how the conflict in Syria, having devasted the healthcare system, has affected Syrian healthcare workers (Joseph et al. 2016). We provide a secondary analysis of a summer 2019 survey Physicians for Human Rights (PHR) conducted with 82 Syrian healthcare workers living in neighbouring countries as well as in northeast and northwest Syria. Our descriptive analysis found that 48 participants reported an average of 16.52 hours of work per day, and 40 participants reported caring for an average of 43 patients per day while working in Syria during the conflict. Sixty-eight participants reported facing barriers to perform their work, and 59 participants reported facing risks as a medical professional. Seventy-one participants experienced traumatic events during their work as a medical professional, and 70 participants experienced stress in the month prior to being interviewed.

This analysis illustrates the negative effect an armed conflict has on healthcare workers through disruptions in their workload, training and resources, barriers and risks faced, and mental health and security issues. The study indicates that these factors require long-term consideration in order to protect healthcare workers and improve the health system.

## I. Introduction/Background

Since 2011, Syria has been engulfed in a complex civil war marked by both targeted and indiscriminate attacks on civilians and civilian infrastructure. The ongoing conflict and resulting humanitarian crisis have left over 5.6 million Syrians as refugees (United Nations High Commissioner for Refugees 2020), 6.2 million internally displaced (United Nations High Commissioner for Refugees), and a documented 380,636 dead by the start of 2020—with the true death toll estimated to be exponentially higher (Ri et al. 2019; Syrian Observatory for Human Rights 2020).

The ongoing armed conflict in Syria has severely impacted the country’s healthcare system. Since the conflict began in 2011, the health sector has suffered from systematic and widespread attacks against healthcare facilities and medical workers (Haar et al. 2018;Physicians for Human Rights 2020). Healthcare professionals in Syria have been directly targeted and killed, as well as systematically persecuted through legalized and extra-judicial means, including forced disappearance, detention, torture, and execution (Fouad et al. 2017). Physicians for Human Rights (PHR), a U.S.-based international advocacy organization, has documented the killing of 923 medical professionals since 2011 (Physicians for Human Rights 2020). In addition to these deaths, Syrian healthcare workers suffer from frequent threats, the destruction of health infrastructure, lack of medical supplies and resources, and lack of surveillance and monitoring capacity (Abbara et al. 2020; Assistance Coordination Unit 2018; Haar et al. 2018). All of these factors, along with the complexities of working in a dynamic and insecure context, have affected the ability of Syrian health professionals to treat their patients.

There is mounting evidence regarding the severe health impacts of the Syrian conflict on the population at large, including the rise of infectious diseases (Abbara et al. 2020; Assistance Coordination Unit 2015; Taleb et al. 2015), non-communicable diseases (Abbara et al. 2015), and mental health issues (Jefee-Bahloul et al. 2015). Past research has also examined the conflict’s effects on medical workers specifically in besieged areas as well as attacks on healthcare (Fardousi et al. 2019; Fouad et al. 2017; Koteiche et al. 2019). The contributions of past research have demonstrated a heavily deteriorated health sector with a significant impact on the ability and willingness of healthcare workers to continue practicing their profession. Due to the restrictions and safety concerns that exist in this climate, there is a critical need for information about the experiences of Syrian healthcare workers themselves and their perspectives on how the conflict has impacted them and their work.

This study aims to explore how the conflict has affected Syrian healthcare workers and their ability to provide their services. In particular, we aim to elucidate (1) the workload, training, and resources of Syrian healthcare workers during the conflict from 2011 to 2019; (2) the practical barriers and risks faced by Syrian healthcare workers; and (3) the mental health and security concerns of these workers during the conflict. We hope to gain a greater understanding of these issues through a statistical analysis of survey data collected by PHR from July 2019 to November 2019 of medical professionals currently living in northeast and northwest Syria as well as outside of the country.

## I. Methods

We collected secondary survey data from PHR that was used for another purpose. We compiled the data into an excel sheet and selected 32 of the 46 survey questions relevant to our research question. We did not include questions relating to detention, as these findings were already published in a separate PHR report (Koteiche et al. 2019). After cleaning the data and translating the interview answers that were in Arabic into English, we categorized the data into healthcare worker characteristics and conflict experiences. Healthcare worker characteristics categories include the participants’ type of health specialty, gender, location of origin and work, and type of work setting while they were working in Syria. The questionnaire included open-ended questions; therefore, we categorized the responses based on an inductive analysis of the concepts and themes that emerged. We utilized a deductive approach for specific frequently used terms, i.e. when terms were used in four or more participant’s answers, that theme became a subcategory. Conflict experiences were categorized into three categories: 1) healthcare worker workload, training and resources, 2) their barriers and risks faced, and 3) their personal health and security. See appendix for definitions of each category and subsequent subcategory.

### Inclusion and Exclusion Criteria

The selection criteria for participants included being a Syrian national and having worked as a healthcare worker (HCW) in Syria during the conflict (2011-2019).Healthcare workers were defined as those professionally involved in the search for,collection, transportation, diagnosis or treatment – including first aid treatment – of the wounded and sick, and in the prevention of disease. Healthcare workers could include physicians, nurses, paramedics, ambulance drivers, search and rescue personnel, and others. The exclusion criteria included being under the age of 18;being unable to provide consent due to linguistic barriers, cognitive impairment, or other disability; or having been engaged in military combat at the time of targeting for the interview.

### Sampling and Data Collection

PHR surveyed Syrian healthcare workers from July 2019 to November 2019,initially to identify healthcare workers who had been detained during the conflict (Koteiche et al. 2019). Through PHR’s network, members of the research team who were connected to displaced Syrian healthcare workers invited them to participate in the study. Those who gave their informed consent participated in the initial surveys. At the end of each survey, the participants were asked to recommend another potential participant for the study (snowball sampling). Survey participants were based in Turkey, Lebanon, Jordan and northwest and northeast Syria. Though all were based out of areas outside of Syrian government control at the time of this survey, participants had previously resided and worked in opposition, government and/or ISIS-controlled areas.

### Survey Design

PHR’s survey sought to collect the following information: demographic;biographical; professional; trauma-related; and detention-related, and data about their professional and personal experiences in Syria during the conflict as well as information on their mental health. Questions were asked in a variety of formats, including yes/no, Likert scale, checklist, and open-ended. (See survey draft in appendix for specific questions). The questionnaire was translated from English into Arabic and then back-translated. The survey questionnaire consisted of a total of 46 questions and was conducted via phone, with the exception of some surveys in Jordan that were conducted in person. The survey was administered verbally (phone or in-person) and data noted on paper forms, then later entered onto an encrypted, secure platform (Kobo) by PHR researchers.

### Data Analysis

We compiled the data to determine the total responses for each relevant interview question and the numerical count and percent for each category and subcategory of healthcare worker characteristics and conflict experiences as well as the minimum,maximum, and mean for any numerical categories. We also graphed the data into bar graphs to give a visualization and comparison of each subcategory count. All statistical analysis was conducted using R.

### Ethical Considerations

PHR interviewed Syrian healthcare workers residing in northeast and northwest Syria, as well as outside of the country, because these populations are less exposed to danger and risk of reprisals than those remaining within areas controlled by the Syrian government. The researchers minimized the risks by obtaining oral informed consent from survey respondents and recording no identifying data. Researchers also took all necessary precautions to ensure confidentiality of records and of interview sites, subjects, and times. PHR’s Ethical Review Board (ERB) approved this research.

## II. Results

PHR interviewed a total of 82 Syrian healthcare workers. We included data from 71 of the 82 participants. Eleven reported having been directly involved in armed conflict and were excluded from further analysis because the research team could not contextualize what that involvement meant or how it could potentially bias the findings. We utilized data for 32 (out of 46) of the interview questions as they were relevant to the research question. For these questions, an average of 65 participants answered each question, with a minimum of 41 and a maximum of 71.

### Healthcare Worker Characteristics

Of the 71 participants, there were 29 (40.85%) doctors, 13 (18.31%) nurses, eight (11.27%) pharmacists, seven (9.86%) paramedics, and 14 (19.72%) other types of healthcare workers. There were 62 (87.32%) men and nine (12.68%) women among the participants. Of the 69 participants who provided that information, 33 (47.83%)worked in field hospitals, 33 (47.83%) worked in hospitals, 15 (21.74%) worked in clinics, nine (13.04%) worked in humanitarian organizations, and 17 (24.64%) worked in other types of health-related work settings. During the time participants were interviewed, 24 participants remained working as healthcare workers, while the other 47 participants were either unemployed or worked outside of healthcare. The three main governorate locations of origin of the 71 participants include Damascus (35.21%), Daraa (15.49%), and Aleppo (15.49%). The three main governorate locations of work of 67 participants include Damascus (37.31%), Idleb (16.42%), and Daraa (10.45%). Table 1 provides information on healthcare worker characteristics.

**Table 1.** Healthcare Worker Characteristics.

| HCW Characteristic | Category | Count | Percent |
| --- | --- | --- | --- |
| Type of Health Professional Specialty | Doctor | 29 | 40.85 |
|  | Other | 14 | 19.72 |
|  | Nurse | 13 | 18.31 |
|  | Pharmacist | 8 | 11.27 |
|  | Paramedic | 7 | 9.86 |
| **Gender** | Male | 62 | 87.32 |
|  | Female | 9 | 12.68 |
| **Type of Work Setting** | Field Hospital | 33 | 47.83 |
|  | Hospital | 33 | 47.83 |
|  | Other | 17 | 24.64 |
|  | Clinic | 15 | 21.74 |
|  | Humanitarian Org | 9 | 13.04 |

### Conflict Experience

#### Workload, Training and Resources Subcategories

Forty-eight participants reported an average of 16.52 hours of work per day while they were health workers in Syria. Forty participants reported tending to an average of 43 patients per day, with 68.1% being direct patient care. Forty-four (80%) of 55 participants had other healthcare workers with their specialty in their work location and 32 (72.73%) of 40 participants reported that they treated war wounds. Thirty (42.86%) of 70 participants reported they were not trained to conduct the work they performed and 34 (61.43%) of 70 participants did not have the appropriate resources at their disposal to perform their work.

#### Barriers and Risks Subcategories

Participants were asked in an open-ended question what the three main barriers were to perform their work, of which 68 participants responded. As previously stated, when terms were used in four or more participant’s answers, that theme became a subcategory. This open-ended question had five subcategories: Limited Staff and Resources, Targeted, Bombardment, Insecurity, and Violence. (See appendix for definitions of each subcategory). Under the ‘Staff and Resources’ subcategory, 33 (48.53%) of 68 participants reported having limited medical supplies, 23 (33.83%) reported having limited qualified specialists, five (7.35%) reported having no funding, and three (4.41%) reported having inadequate health facilities, no training,and flight of healthcare workers each. Under the ‘Targeted’ subcategory, 13 of 68 participants (19.12%) experienced attacks on health facilities and 12 (17.65%) were directly targeted. Under the ‘Bombardment’ subcategory, 23 of 68 participants (33.82%) experienced civilian bombardment and blockades and eight (11.76%)experienced limited transportation of medical supplies and patients. Under the ‘Insecurity’ and ‘Violence’ subcategories, 26 of 68 participants (38.24%) felt unsafe and 10 (14.71%) experienced violence respectively (International Committee of Red Cross). (See Figure 1).

**Figure 1:**
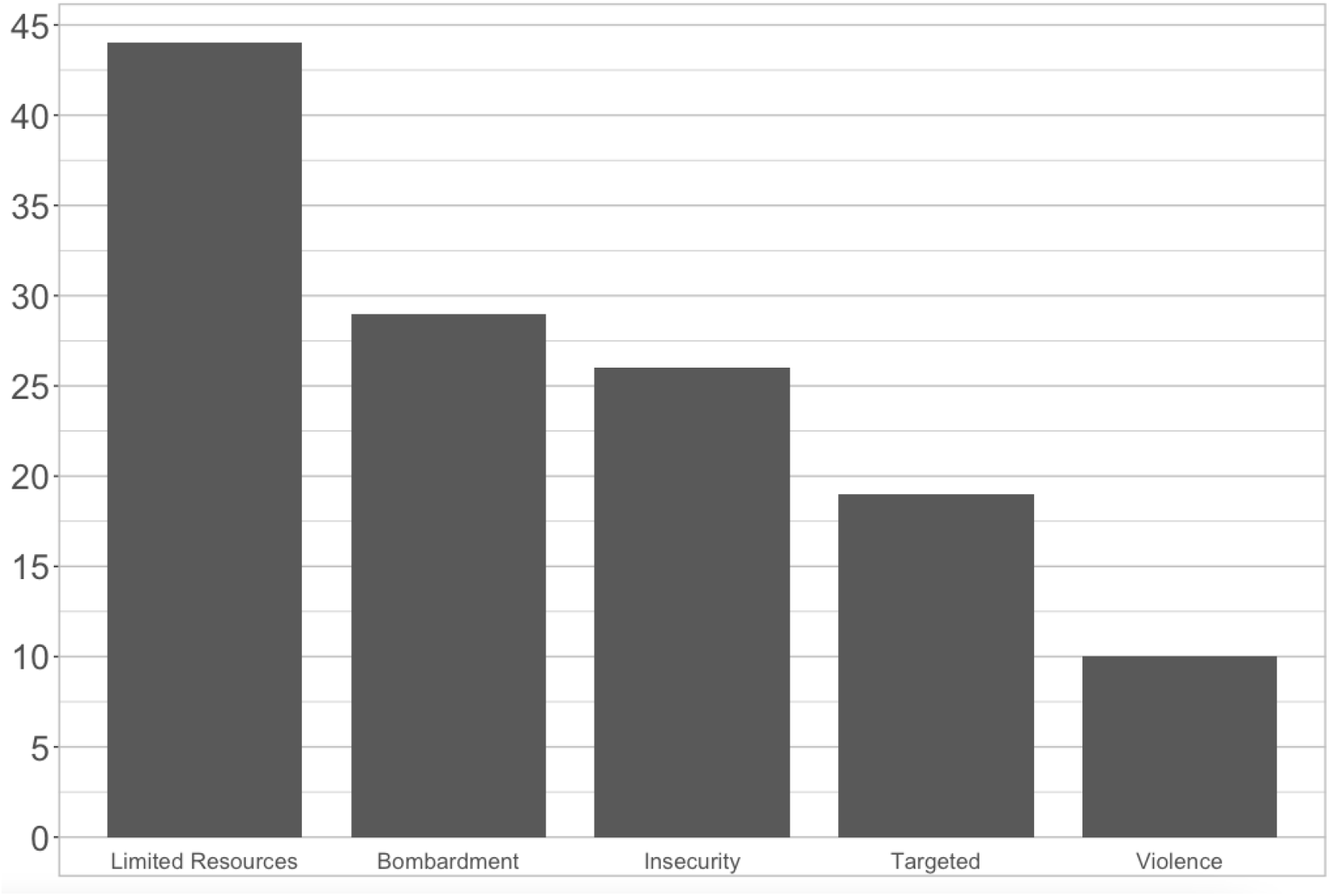
HCW Barriers faced in performing their work.

Participants were asked in an open-ended question what the three main risks were as a medical professional, of which 59 participants responded. This open-ended question had four subcategories: Bombardment, Detention, Insecurity, and Targeted. (See appendix for definitions of each subcategory). Under the ‘Bombardment’ subcategory, 25 (42.37%) of 59 participants experienced civilian bombardment and blockades. Under the ‘Detention’ and ‘Insecurity’ subcategories, 21 (35.59%) were detained or arrested and 17 (28.81%) felt unsafe respectively. Under the ‘Targeted’ subcategory, 15 (25.42%) faced death threats, 10 (16.95%) were directly targeted, and nine (15.25%) experienced attacks on health facilities.

#### Mental Health and Security Subcategories

Sixty-five participants were forcibly displaced an average of three times, with 16 (24.62%) displaced once, 21 (32.31%) displaced twice, 15 (23.08%) displaced three times, nine (13.85%) displaced four times, and four (6.15%) displaced five times.

Participants were asked in an open-ended question what their reasons were for departing Syria from where they worked as healthcare workers, of which 66 participants responded. From this open-ended question, six subcategories emerged: Insecurity, Violence, Bombardment, Detention, Targeted, and Recapture of Area. (See appendix for definitions of each subcategory). Under the ‘Insecurity’ subcategory, 24 (36.36%) of 66 participants left due to feelings of unsafety, including fear for themselves and their family. Under the ‘Violence’ subcategory, 13 (19.70%) left due to armed forces, shootings, the participants’ desertion of the army and their fear of being drafted. Under the ‘Bombardment’ and ‘Detention’ subcategories (22.73%) of the 66 participants that departed from Syria left due to civilian bombardment and blockades and 10 (15.15%) left due to past detention or arrest respectively. Under the ‘Targeted’ and ‘Recapture of Area’ subcategories, 16 (24.24%) left due to being wanted by the Syrian government and fear of being arrested and six (9.09%) left due to the Syrian government’s recapture of the area the participant worked respectively.

Participants were asked whether they resided as practicing healthcare workers in Syrian government-controlled, opposition-controlled, or ISIS controlled areas before they departed Syria. Of the 67 participants that responded, 22 (32.84%) resided in Syrian government-controlled areas, 39 (58.21%) resided in opposition-controlled areas, and eight (11.94%) in ISIS controlled areas.

Participants were asked a series of yes/no questions regarding experiencing traumatic events during their work as healthcare workers, in which all 71 participants (100%) responded. Of these, 66 (92.96%) said yes to facing attacks including direct bombardments, battles, or other types of large-scale violence, 65 (92.86%) said yes to feeling that their life was threatened because they were working as a medical professional in Syria, 50 (71.43%) said yes to personally being threatened with death or injury, 52 (73.24%) said yes to feeling intense fear, helplessness, or horror and 22 (31%) said yes to being physically injured. (See Figure 2).

**Figure 2:**
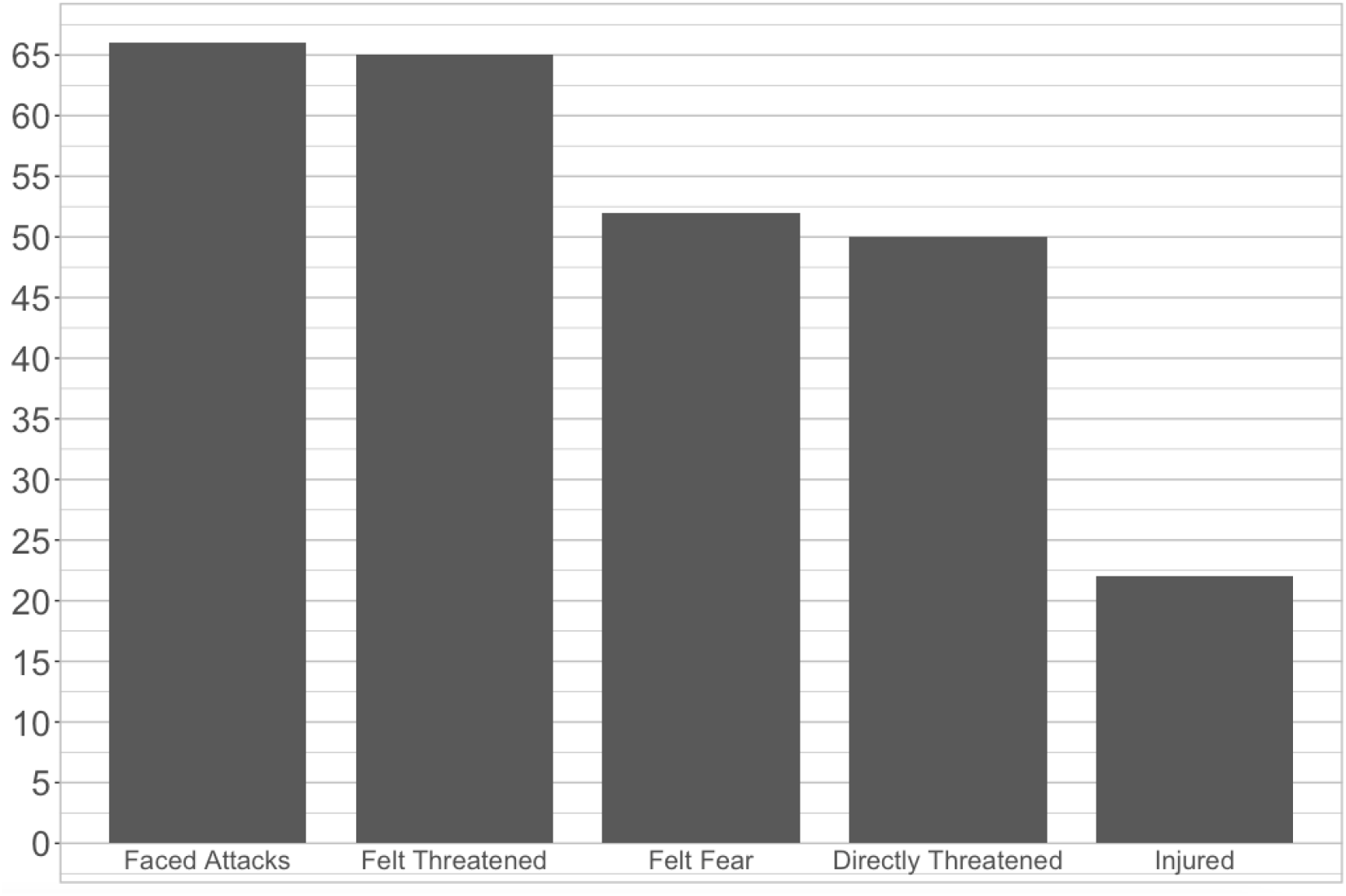
Medical professionals that experienced traumatic events during their work.

Participants were asked a series of yes/no questions regarding experiencing stress over the past month when interviewed, of which 70 participants responded. Of these, 51 (72.86%) said yes to trying hard not to think about traumatic events or avoiding situations that reminded them of the events, 42 (60%) said yes to having nightmares about the events, 40 (57.14%) said yes to being on constant guard, watchful, or easily startled, 36 (52.17%) said yes to feeling numb or detached from people, activities, or their surroundings, and 31 (44.29%) said yes to feeling guilty. (See Figure 3).

**Figure 3:**
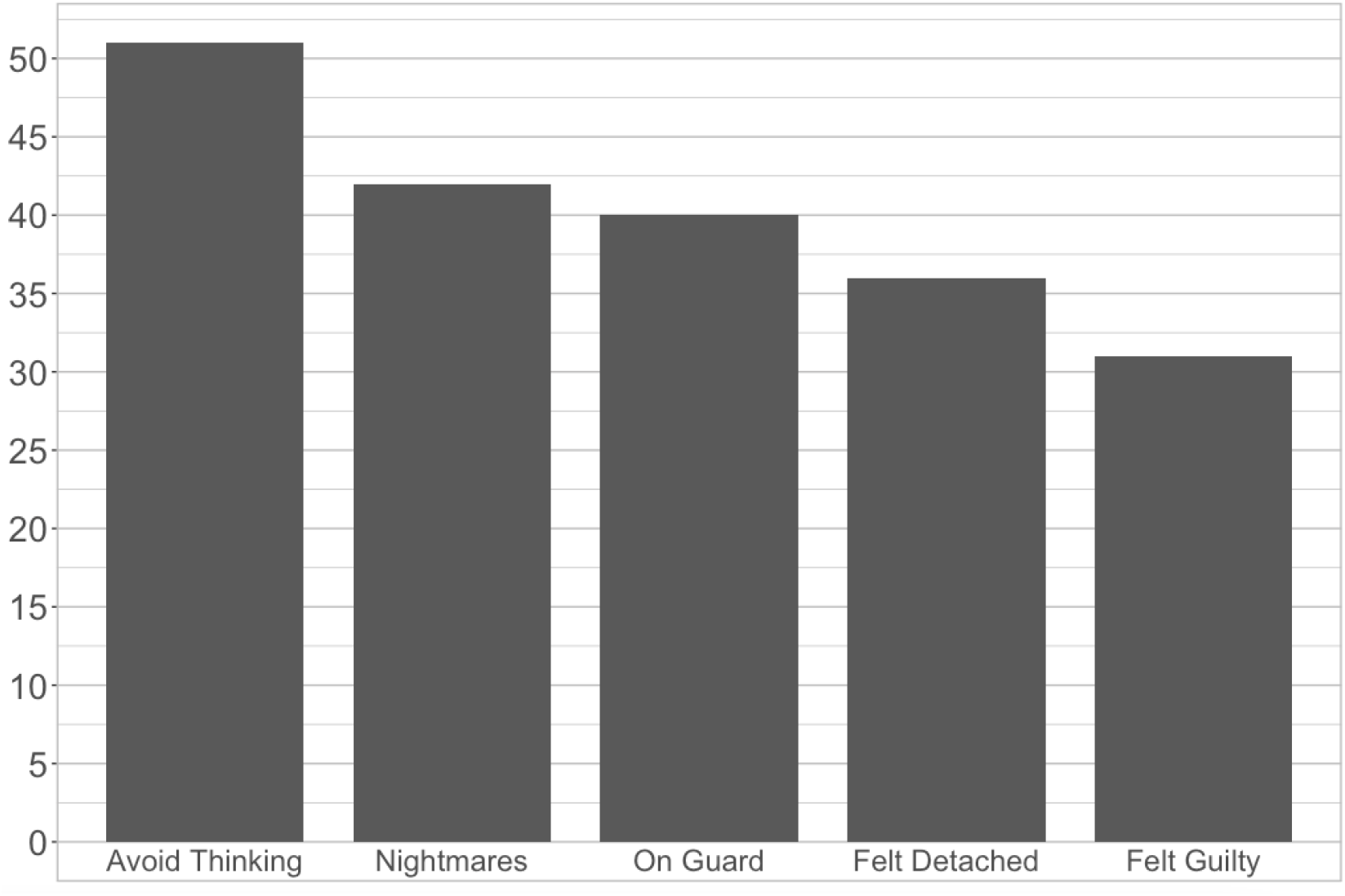
Health professionals that experienced stress over the past month.

## Discussion

This study provides a deeper understanding of healthcare worker experiences during conflict through a detailed descriptive analysis exploring healthcare workers’ heavy workloads, barriers and violence they faced and the impact of their experiences on their mental health. To our knowledge, these interviews with 24 current and 47 former Syrian healthcare workers represent the largest sample of healthcare workers in Syria to report on their experiences.

We found that 48 participants reported an average of 16.52 hours of work per day and 40 participants reported tending to an average of 43 patients per day, with 68.1%being direct patient care. Comparing this to physicians in the US that work an average of 51 hours per week (10 hours per day) and tend to an average of 20 patients per day (Elflein 2018) illustrates the severe work burdens of these healthcare workers. Limited medical supplies, attacks on health facilities and limited transportation of medical supplies and patients lead to many healthcare workers not having the appropriate resources to treat their patients, exacerbating the burden. Many of the healthcare workers also reported they were not trained to perform their work, where most had to treat war wounds due to civilian bombardment and violence, suggesting that task shifting was not concomitant with appropriate training.

All the participants interviewed experienced traumatic events during their work as a medical professional including being personally threatened with death or injury, feeling intense fear, helplessness, or horror, experiencing the loss of colleagues or family members and being physically injured. Those that were directly targeted and threatened because they were healthcare workers and were detained or arrested felt unsafe and departed from their place of residence, many of which were displaced more than once. Their experiences working as medical professionals during a conflict have impacted their mental health, with 70 of 71 participants reporting having experienced nightmares and avoiding situations that reminded them of their experience, in addition to experiencing feelings of guilt.

### Limitations

There are several important limitations to this study. Although larger than other studies of healthcare workers in conflict, the sample size was small. Participants were selected through PHR’s network and secondary snowball sampling; thus, the sampling was not random and raises the risk of selection bias. The sample was also not representative of the whole population, including the geographic distribution of participants and the majority of participants being men. Furthermore, the data used in this study was secondary and the interviews were initially conducted to identify respondents who had specific experiences of detention or targeting which limits the number and type of questions asked. While the study provides insights into the experiences of healthcare workers, we are not able to probe deeper into how or why conflict experiences resulted in these responses. Further qualitative work could explore how conflict, or attacks on health specifically, impacted healthcare workers and more randomized or community based quantitative work may help avoid the biases associated with the heterogeneity of this dataset.

## Conclusion

This study explores how the conflict in Syria has had a negative effect on healthcare workers through their workload, training and resources, barriers and risks faced, and mental health and security. The flight of medical professionals, attacks on health facilities, bombardment of civilians and civilian structures, and overall violence led to many healthcare workers being overworked while managing with little to no training and limited medical resources to perform the work they are tasked with, all of which impacts their mental health. Without additional and meaningful resources, training,and protection, healthcare workers will continue to face enormous stress, high work burdens and mental health consequences that ultimately weaken the entire health system.

Healthcare workers in conflict settings are crucial functioning civil society and have a critical role in providing care to all people, regardless of political affiliation or other factors. Now after 9 years of brutal conflict, the Syrian government as well as international stakeholders must adopt measures to combat the severe shortage and prevent continued flight of medical professionals. This can be done by providing adequate training and salaries to all healthcare workers as well as ensuring their safety regardless of their location, affiliation of medical facilities where they work, political affiliations, or the civilian populations they serve. The Syrian government and international community should also maintain an equal and adequate supply chain of resources including medical equipment, medication, vaccines, and safe transportation of medical supplies and patients. This includes ensuring the successful provision of humanitarian aid deliveries to populations living outside of government-controlled areas and the hospitals that serve them through all available avenues,including by restoring essential cross-border aid mechanisms through both the Northeast and Northwest. In recently ‘reconciled’ areas, such as Eastern Ghouta and Daraa, which have changed from opposition to government control, the Syrian government needs to provide safety and access to work for medical professionals residing there and ensure that they are not subject to retaliation for their work. The international community should hold the Syrian government accountable for protecting healthcare workers and health facilities.

The systematic attacks on civilians and civilian infrastructure and the targeted attacks of the health sector and other violations of international humanitarian law have not only contributed to the flight of medical personnel out of Syria; they have severely eroded the professional capacities and the mental and physical health of those who have remained within the country. The effect of this on the health system and on the communities, they serve has been catastrophic.

## Supporting information

Cover Letter

## Data Availability

Data noted on paper forms, then later entered onto an encrypted, secure platform (Kobo) by PHR researchers

## Acknowledgments

We thank Rayan Koteiche and other staff at Physicians for Human Rights for conducting this unique healthcare worker survey and allowing us to use their data. We thank Joseph Leone for assisting us on the manuscript writing. Finally, we thank all the healthcare workers that participated in the questionnaire.

## Appendix

### Definition of Subcategories

*Workload, Training and Resources Subcategories:*

1. *‘Training and Resources’* including:
  a. *‘Other HCWs with your specialty’* referred to the number of participants that had other medical professionals with the participant’s specialty/skill set in their location.
  b. *‘Treated wounds’* referred to the number of participants that treated war wounds.
  c. *‘Trained to Conduct Work’* referred to the number of participants that were trained to conduct that type of work/provide that type of treatment.
  d. *‘Resources’* referred to the number of participants that felt they had the appropriate resources at their disposal to perform their work.
2. *‘Hours/Day’* referred to the number of hours a day the participant worked on average.
3. *‘Percent Care’* referred to percent of time that was dedicated to providing direct patient care.
4. *‘Nb Patients/Day’* referred to the average number of patients a day the participant tended to.

*Barriers and Risks Subcategories:*

1. *‘Barriers’* referred to the number of participants that faced barriers to performing their work, where the barriers included: *(Open-ended question)*
  a. *‘Limited Staff and Resources’* which referred to limited medical supplies (medication, equipment, resources), qualified specialists and training, inadequate health facilities, flight of HCWs, and no funding.
  b. *‘Bombardment’*: which referred to civilian bombardment and blockades and limited transportation of medical supplies and patients.
  c. *‘Insecurity’* which referred to feelings of unsafety.
  d. ‘*Targeted*’: which referred to HCWs being targeted, attacks on health facilities.
  e. ‘*Violence*’ which referred to armed forces, shootings, the participants’ desertion of the army and their fear of being drafted.
2. *‘Risks’* referred to the number of participants that faced risks as a medical professional, where the risks included: *(Open-ended question)*
  a. *‘Targeted’* which referred to HCWs being targeted, attacks on health facilities, and death threats.
  b. *‘Bombardment’* which referred to civilian bombardment and blockades.
  c. *‘Insecurity’* which referred to feelings of unsafety and random kidnappings.
  d. *Detention’* which referred to being detained or arrested.

*Personal Health and Security subcategories:*

1. *‘Amount of Times Displaced’* referred to the number of participants that were displaced categorized into numerical categories.
2. *‘Departure Reason’* referred to the number of participants whose reason for departure included: (Open-ended question)
  a. ‘*Insecurity’* which referred to feelings of unsafety and fear for themselves and their family.
  b. *‘Violence’* which referred to armed forces, shootings, the participants’ desertion of the army and their fear of being drafted.
  c. *‘Bombardment’* which referred to civilian bombardment and blockades.
  d. *‘Detention’* which referred to being detained or arrested.
  e. *‘Targeted’* which referred to being wanted by the Syrian government and fear of being arrested.
  f. *‘Recapture of Area’* which referred to the Syrian government’s recapture of the area the participant worked.
3. *‘Control of Area’* referred to the number of participants that worked in an area controlled by the Syrian government, opposition groups or ISIS.
4. *‘Traumatic Events’* referred to the number of participants during their work as a medical professional that experienced:
  a. *‘Attacks’* which referred to direct bombardments, battles, or other types of large-scale violence.
  b. *‘Felt Threatened’* which referred to feeling that their life was threatened because they were working as a medical professional in Syria.
  c. *‘Felt Fear’* which referred to feelings of intense fear, helplessness, or horror.
  d. *‘Threatened’* which referred to personally threatened with death or injury.
  e. ‘Injured’ which referred to being physically injured
5. *‘Stress Indicators’* referred to the number of participants that experienced stress over the past month including:
  a. *‘Avoid Thinking’* which referred to trying hard not to think about traumatic events or avoiding situations that reminded them of the events.
  b. *‘Nightmares’* which referred to having nightmares about the events or thinking of the events when they did not want to.
  c. *‘On Guard’* which referred to being constantly on guard, watchful, or easily startled.
  d. *‘Felt Detached’* which referred to feeling numb or detached from people, activities, or your surroundings.
  e. *‘Felt Guilty’* which referred to feeling guilty or unable to stop blaming themselves or others for the events.

